# Cultural Adaptation of a Brief Intervention to Reduce Alcohol Use Among Injury Patients in Tanzania

**DOI:** 10.1101/2021.07.21.21260811

**Authors:** Armand Zimmerman, Msafiri Pesambili, Ashley J. Phillips, Judith Boshe, Blandina T. Mmbaga, Michael H. Pantalon, Monica Swahn, Jon Mark Hirshon, Joao Ricardo Nickenig Vissoci, Catherine A. Staton

## Abstract

**Background:** Harmful alcohol use is a leading risk factor for injury-related death and disability in low- and middle-income countries (LMICs). Brief negotiational interventions (BNIs) administered in emergency departments (EDs) to injury patients with alcohol use disorders (AUDs) are effective in reducing post-hospital alcohol intake and re-injury rates. However, most BNIs to date have been developed and implemented in high-income countries. The efficacy of BNIs in LMICs is largely unknown as few studies have undertaken the rigorous task of culturally adapting these interventions to new settings. Given the high prevalence of alcohol-related injury in the Kilimanjaro region of Tanzania, we culturally adapted a BNI to reduce post-injury alcohol use for implementation in this patient population.

**Methods:** We used an iterative, multiphase process to culturally adapt a high-income country standard of care BNI to the Tanzanian setting using the Intervention Mapping ADAPT framework. Our team consisted of local healthcare professionals with extensive experience in counseling patients who use alcohol, as well as an international team of academic and clinical professionals. Focus groups were used to inform culturally appropriate changes to the standard of care BNI protocol. Objective assessment of BNI delivery was performed to ensure adherence to the FRAMES model of motivational interviewing.

**Results:** We developed the Punguza Pombe Kwa Afya Yako (PPKAY); a one-time, 15-minute nurse-led BNI that encourages safe alcohol use and motivates change in alcohol use behaviors among injury patients in the Kilimanjaro region of Tanzania. Adaptations to the original intervention protocol include changes regarding the interventionist, how a patient is greeted, how the topic of alcohol use is raised, how a patient is informed of their harmful alcohol use, how graphics are visualized within the intervention protocol, how behavior change is motivated, and which behavior changes are encouraged.

**Conclusions:** The PPKAY intervention is the first BNI to be culturally adapted for delivery to injury patients in an LMIC population. Our study demonstrates a unique approach to adapting substance use interventions for use in LMICs, and shows that cultural adaptation of alcohol use interventions is feasible even in settings where community knowledge regarding harmful alcohol use is limited. Our study prompts the need for further research and cultural adaptation of BNIs for other low-income communities at increased risk of alcohol-related harm.

## INTRODUCTION

Globally, harmful alcohol use causes an estimated 3 million deaths each year, resulting in an annual mortality rate that exceeds that of HIV/AIDS, malaria, and tuberculosis (WHO, 2018). Similarly, injuries account for 5.8 million of the world’s total annual deaths, making it a leading contributor to global mortality (WHO, 2014). Among all deaths resulting from harmful alcohol use, 28.7% are caused by injuries (WHO, 2018).

Low- and middle-income countries (LMICs), specifically those in Africa, carry the greatest burden of both alcohol consumption and injury rates (WHO, 2018; WHO, 2014; Haagsma et al., 2016). The global rate of injury deaths per 100,000 people is 80.1 (WHO, 2014). However, this rate increases to 98.8 per 100,000 people in the World Health Organization (WHO) African region (WHO, 2014). Similarly, the global rate of alcohol consumption is 32.8 grams of pure alcohol per day per drinker whereas in Africa this rate increases to 40.0 grams of pure alcohol per day per drinker (WHO, 2018). Moreover, alcohol consumption is a leading risk factor for morbidity among 15-24 year-old males in sub-Saharan Africa and is only expected to increase in coming years (Gore et al., 2011; Ferreira-Borges et al., 2017).

Within sub-Saharan Africa, Tanzania has high rates of alcohol-related injury. At Muhimbili National Hospital in Dar es Salaam, it is estimated that 46.9% of patients presenting with injuries to the emergency department (ED) test positive for alcohol (Mundenga et al., 2019). Similarly, at a large referral hospital in the Kilimanjaro region, it is estimated that 30.0% of injury patients presenting to the ED test positive for alcohol (Staton et al., 2018). In addition, the prevalence of alcohol use disorders (AUDs) in Tanzania is high. In a survey of 1954 men and women aged 15-24 years from Kilimanjaro and Mwanza regions, 10.5% screened positive for AUD (Francis et al., 2015). In a similar survey of 899 men and women aged 18-59 years from Dar es Salaam, the prevalence of hazardous drinking was 5.7% (Mbatia et al., 2009). The most recent WHO estimate places the prevalence of AUD in Tanzania at 6.8% (WHO, 2018). Harmful alcohol use appears to be higher among injury patients presenting to EDs when compared to Tanzania’s general population. The prevalence of AUD among injury patients may therefore be higher than what is observed across the general population. Consequently, alcohol use reduction interventions targeting ED injury patients may be an effective and efficient means of reducing overall harmful drinking and injury rates in the population.

A brief negotiational intervention (BNI) is a single session interview, lasting no more than 30 minutes, which attempts to reduce harmful post-hospitalization alcohol use (Meredith S.H. Landy et al., 2016). In high- and middle-income countries, the delivery of BNIs in the emergency or other departments has been proven effective in reducing both post-hospital alcohol consumption and post-hospital alcohol-related injury (Meredith S.H. Landy et al., 2016; Mcqueen et al., 2015; Joseph et al., 2017; Baldacchino et al., 2018). However, the efficacy of a BNI in reducing harmful alcohol use among injury patients in Tanzania is unknown. Studies assessing the impact of BNIs on post-hospital alcohol consumption in Africa have occurred predominantly in South Africa with a focus on non-injury patients (Sorsdahl et al., 2017; Peltzer et al., 2013; Pengpid et al., 2013; Huis In ’t Veld et al., 2012; Peltzer et al., 2008). To optimize the effectiveness of a BNI in producing a desired outcome, the BNI must be sensitive to a patient’s culture and environment. Patient’s in different regions of the world may experience vastly different stressors that influence harmful alcohol and which result from unique cultural norms or acculturation experiences. The cultural adaptation of BNIs to account for such stressors among subpopulations with specific demographics is shown to be effective in improving patient outcomes (Satre et al., 2015; Field et al., 2015).

The aim of this study was to develop a BNI culturally adapted for delivery to injury patients presenting to Kilimanjaro Christian Medical Centre (KCMC) in Moshi, Tanzania. Our objective in designing the BNI was to reduce post-hospitalization alcohol consumption and therefore re-injury rates among those receiving the intervention.

## METHODS

### Setting

KCMC is a zonal tertiary referral hospital serving a population of 15 million people in Northern Tanzania (KCMC Website). Over 2000 injury patients present to the KCMC ED each year, or an average of about 46 injury patients per week (Staton et al., 2018). The prevalence of day of injury alcohol use among this injury patient population is 30% (Staton et al., 2018). Moreover, it has been shown that greater alcohol consumption is associated with increased odds of injury among patients presenting to the KCMC ED (Staton et al., 2018). The KCMC ED is therefore an appropriate setting for the administration of a BNI designed to reduce post-injury alcohol use.

### Intervention Mapping

#### ADAPT strategy

To ensure rigor and reproducibility of our adaptation process, we adhered to the central tenets of Intervention Mapping (IM) in planning out our adaptation protocol. IM is a framework for the planning of health promotion programs (Fernandez et al., 2019) and has been used to adapt already existing interventions.(Link ADAPT) The IM framework contains six steps: (1) Needs assessment, organizational capacity, and logic models, (2) Search for evidence-based interventions (EBIs), (3) Judge behavioral and environmental fit and list adaptations (4) Adaptation of materials and activities, (5) Planning of implementation, and (6) Write evaluation questions (add ADAPT PAPER)

This manuscript will briefly present all steps, but will mostly focus on Step 4, the adaptation process in greater detail.

##### Step 1:Needs assessment, organizational capacity, and logic models

###### Needs assessment

For this study, IM steps 1-2 are embodied by the discussions that preceded our research. Given the high prevalence of harmful alcohol use in Tanzania as documented by peer-reviewed literature and our team’s prior work, our ultimate goal was to drive forward efforts to reduce harmful alcohol use in Tanzania (Staton et al., 2018; Staton et al., 2018). Our prior work highlighted limited knowledge about alcohol use and its related harms as well as an external focus of control among patients calling for a motivational theory of behavior change coupled with education for our intervention.

###### Organizational capacity

Our intervention selection was informed by resource availability in Tanzania. Since numerous hospitals across Tanzania are low-resource with regard to both staff and equipment, it made sense to select an easy to teach intervention, such as the BNI, which does not necessitate equipment or significant staff time.

###### Team structure

We assembled a team of individuals who could inform culturally appropriate changes to the model BNI protocol without disrupting the protocol’s fidelity. Our team consisted of local and external professionals. Local professionals included KCMC nurses, KCMC research assistants, and KCMC substance use counsellors all with previous experience working with patients with AUDs. External professionals included clinical psychologists, epidemiologists, and medical doctors from the United States all with extensive experience researching substance abuse. Upon assembling our team we held multiple instructional team meetings to ensure all team members understood the purpose, components, and delivery procedure of the model BNI protocol.

###### Logic model of problem

https://app.diagrams.net/#G1zWhICB9PmjwKKWh-uJT-fLGyRSF_Rlpa

##### Step 2: Search for evidence-based interventions (EBIs)

###### SBIRT

Our initial, or model, BNI was a National Institute on Alcohol Abuse and Alcoholism (NIAAA) funded Screening and Brief Intervention and Referral to Treatment (SBIRT) plan for a United States ED setting. This brief intervention incorporates FRAMES into a four-step protocol (NIAAA Manual). The model BNI was developed to lower the risk of alcohol use among ED patients in the United States. We used this BNI as a starting point for creating a culturally adapted BNI for injury patients in Tanzania.

###### FRAMES

The structure of BNIs is based largely on the Feedback, Responsibility, Advice, Menus, Empathy, and Self-efficacy (FRAMES) model of motivational interviewing developed by Miller and Sanchez (1993) (Bien et al., 1993). The acronym FRAMES comprises six elements shown to optimize the effectiveness of motivational interviews: feedback, responsibility, advice, menu, empathy, self-efficacy (Table 1).

**Table 1.** FRAMES Model

|  |  |
| --- | --- |
| **Feedback** | Give feedback on the risks and negative consequences of alcohol use. Seek the client's reaction and listen. |
| **Responsibility** | Emphasize the individual's responsibility for making his/her own decision about his/her alcohol use. |
| **Advice** | Give straightforward advice on modifying alcohol use. |
| **Menu of options** | Give a menu of options to choose from, fostering a clients involvement in decision making. |
| **Empathy** | Be empathetic, respectful, and non-judgemental. |
| **Self-Efficacy** | Express optimism about his/her ability to modify his/her alcohol use. |

##### Step 3: Assess (detailed) fit and plan adaptations

We assembled and educated our team on the FRAMES model and BNI components through instructional meetings led by the principal investigator. Members were subsequently trained in motivational interviewing by having them observe and practice BNIs. Motivational interviewing practice was facilitated by healthcare professionals at KCMC with previous experience conducting BNIs. In phase two, we had all team members individually identify terminology, phrases, questions, and information within the model BNI that might require adaptation. In phase three, we held focus groups with our team to reach a consensus on what changes to make to the BNI protocol. Focus groups were facilitated by the principal investigator. The principal investigator would ask each team member to present their suggested changes to the BNI protocol. Following each team member’s presentation, all members would engage in an open discussion until a unanimous consensus was reached about whether or not to include the presented changes.

##### Step 4: Adaptations

The goal of Step 4 was to repeat the following sequence of events until our team unanimously agreed that no further adaptations were necessary to deliver the model BNI in the Tanzanian setting: identify areas of the BNI requiring change through focus groups, agree on changes, incorporate changes, observe practice deliveries of the BNI, elicit feedback from all team members. The final adapted BNI protocol and the reasons for our adaptations are presented below. Immediately following each focus group, the BNI protocol was updated accordingly.

Two pairs of team members practiced delivering the new culturally adapted BNI to each other. A designated objective observer assessed delivery of the culturally adapted BNI, from one team member to another, for adherence to the FRAMES model. Each pair practiced delivery until at least 95% adherence to the FRAMES model was obtained on three independent deliveries of the BNI protocol. We then held focus groups with the entire team, including external academic and clinical professionals, to inform final changes to the adapted BNI protocol. Again, these focus groups were facilitated by the principal investigator and used the same format as described above. The results of this manuscript will present this step in greater detail.

##### Step 5: Plan for implementation

While beyond the scope of this manuscript, we started with a feasibility trial and subsequent clinical trial which is ongoing. In this trial we established a protocol for dissemination planning as well as ‘train the trainer’ developments, to create structures for implementation. Circular implementation and evaluation; circular implementation and dissemination

##### Step 6: Plan for evaluation

The evaluation and implementation processes of the ongoing feasibility trial with currently 75 patients are circular and inform each other. The present work was approved by the Kilimanjaro Christian Medical Centre - Tanzania IRB, protocol:Pro00062061.

## RESULTS

For the adaptation process, we created a workflow, presented in Figure 2. We divided the adaptation process into 6 phases: Team Training, Identify Areas for Change, Focus groups to identify cultural and lingual changes to the underlying BI, Practice BI Delivery, Assess BI delivery, Focus groups to finalize the culturally adapted BI.

**Figure 1:**
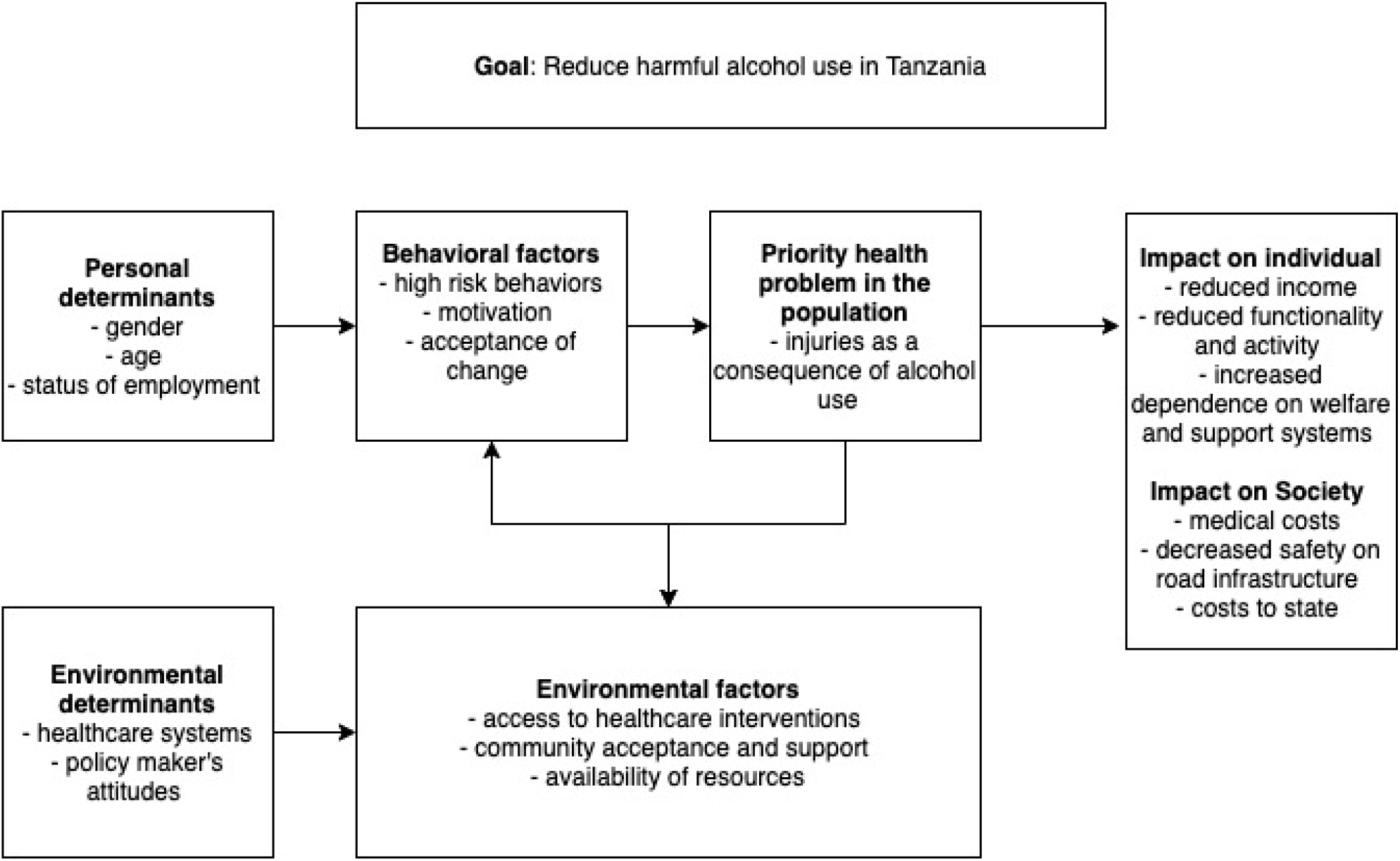
Logic Model of Problem

**Figure 2:**
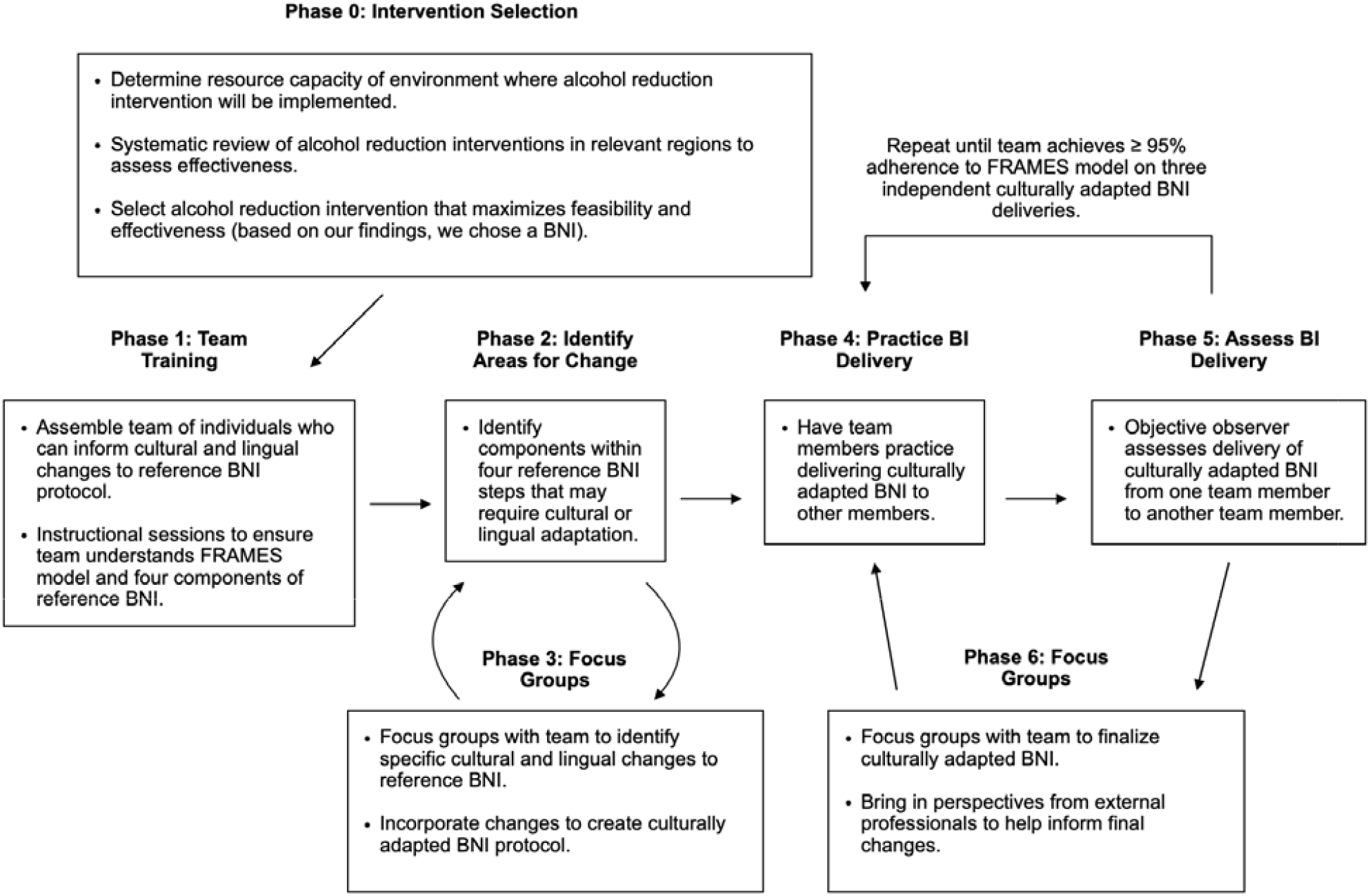
Overview of Adaptation process

### Culturally Adapted BNI Protocol

Using the information gained from our focus groups, we developed the Punguza Pombe Kwa Afya Yako (PPKAY); a one-time, 15 minute, nurse-led motivational interview discussing safe drinking behaviors and which negotiates change in alcohol use for injury patients admitted to the KCMC ED in Moshi, Tanzania (Table 2).

**Table 2.**
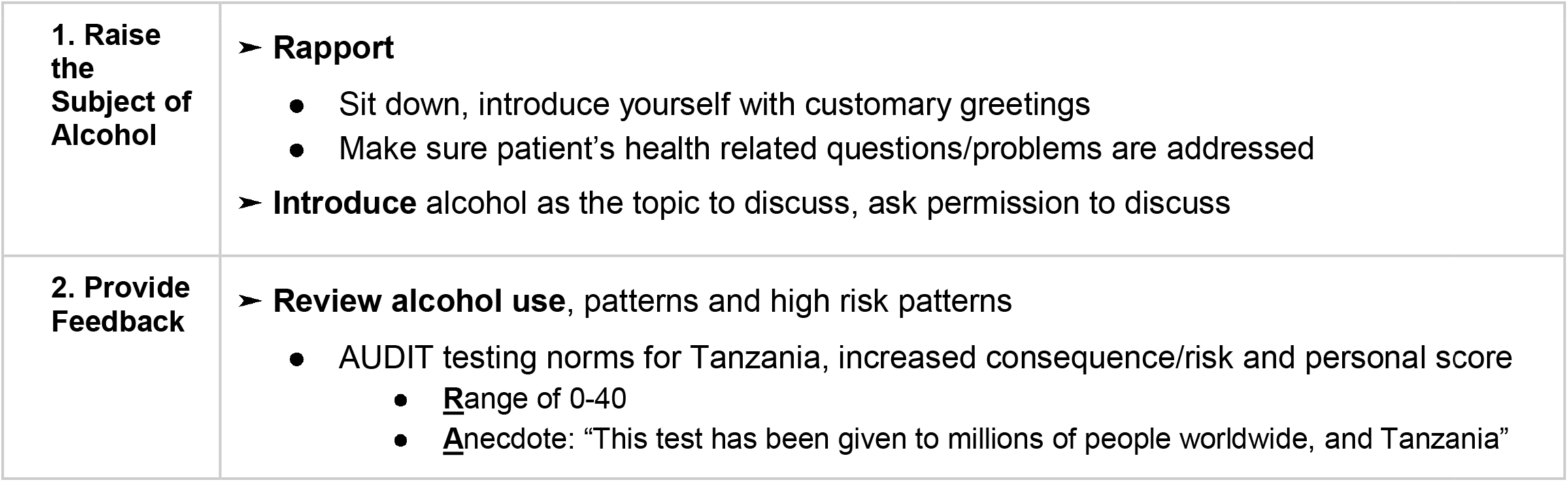
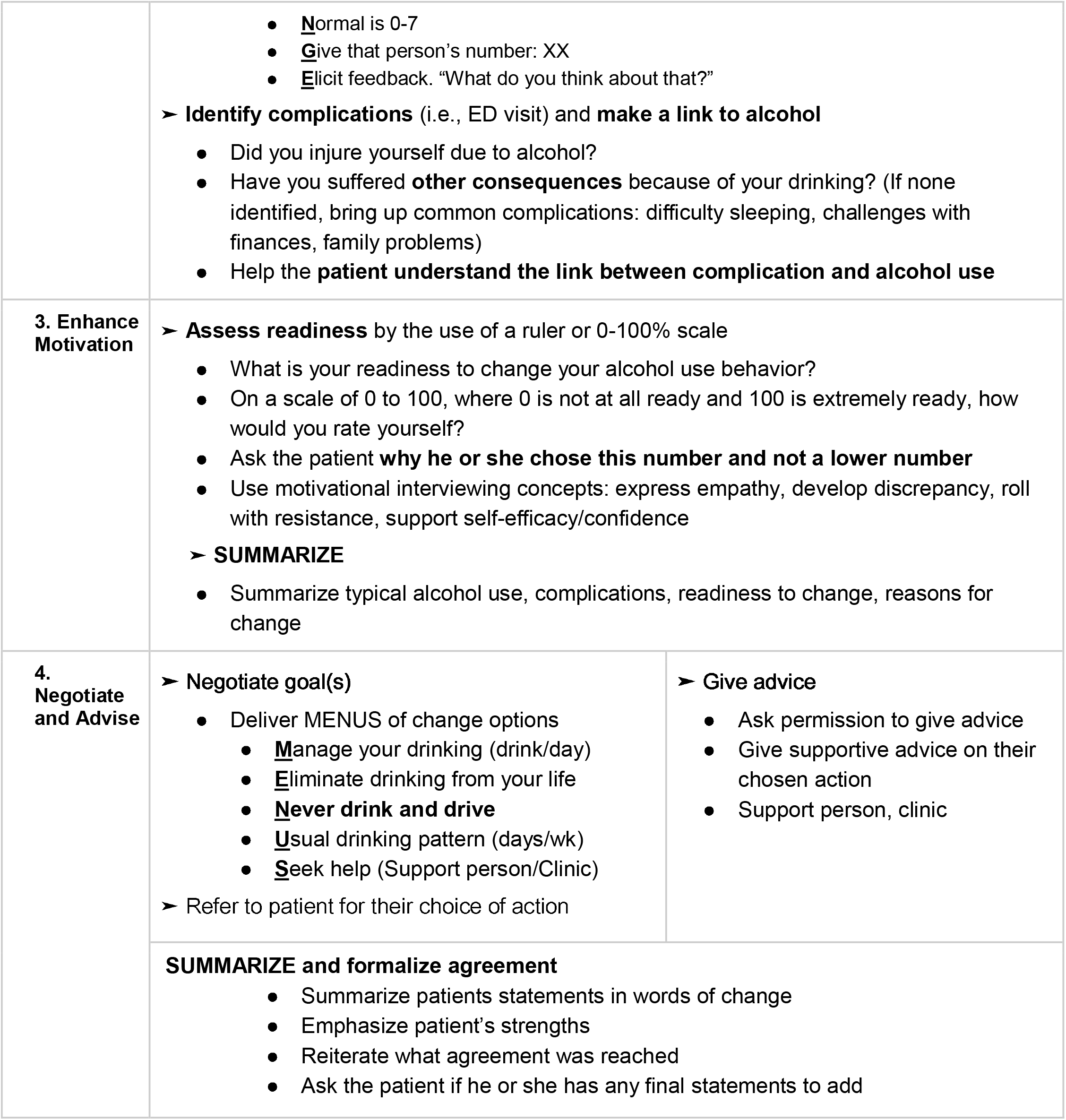
English Version of the PPKAY Protocol.

|  |  |
| --- | --- |
| **1. Raise the Subject of Alcohol** | ➤ **Rapport**<br>• Sit down, introduce yourself with customary greetings<br>• Make sure patient's health related questions/problems are addressed<br>➤ **Introduce** alcohol as the topic to discuss, ask permission to discuss |
| **2. Provide Feedback** | ➤ **Review alcohol use**, patterns and high risk patterns<br>• AUDIT testing norms for Tanzania, increased consequence/risk and personal score<br>• <u>R</u>ange of 0-40<br>• <u>A</u>ncedote: "This test has been given to millions of people worldwide, and Tanzania" |

|  |  |  |
| --- | --- | --- |
|  | • <u>N</u>ormal is 0-7<br>• <u>G</u>ive that person's number: XX<br>• <u>E</u>licit feedback. "What do you think about that?"<br>➤ **Identify complications** (i.e., ED visit) and **make a link to alcohol**<br>• Did you injure yourself due to alcohol?<br>• Have you suffered **other consequences** because of your drinking? (If none identified, bring up common complications: difficulty sleeping, challenges with finances, family problems)<br>• Help the **patient understand the link between complication and alcohol use** |  |
| **3. Enhance Motivation** | ➤ **Assess readiness** by the use of a ruler or 0-100% scale<br>• What is your readiness to change your alcohol use behavior?<br>• On a scale of 0 to 100, where 0 is not at all ready and 100 is extremely ready, how would you rate yourself?<br>• Ask the patient **why he or she chose this number and not a lower number**<br>• Use motivational interviewing concepts: express empathy, develop discrepancy, roll with resistance, support self-efficacy/confidence<br>➤ **SUMMARIZE**<br>• Summarize typical alcohol use, complications, readiness to change, reasons for change |  |
| **4. Negotiate and Advise** | ➤ **Negotiate goal(s)**<br>• Deliver MENUS of change options<br>• <u>M</u>anage your drinking (drink/day)<br>• <u>E</u>liminate drinking from your life<br>• **<u>N</u>ever drink and drive**<br>• <u>U</u>sual drinking pattern (days/wk)<br>• <u>S</u>eek help (Support person/Clinic)<br>➤ Refer to patient for their choice of action | ➤ **Give advice**<br>• Ask permission to give advice<br>• Give supportive advice on their chosen action<br>• Support person, clinic |
|  | **SUMMARIZE and formalize agreement**<br>• Summarize patients statements in words of change<br>• Emphasize patient's strengths<br>• Reiterate what agreement was reached<br>• Ask the patient if he or she has any final statements to add |  |

#### 1: Raise the Subject of Alcohol

The first step of the model BNI protocol establishes rapport with and introduces the topic of alcohol to the patient. Cultural adaptation of step one focused on 1) culturally appropriate ways to establish rapport and open the stage for a motivational discussion, 2) the most appropriate way to introduce the topic of alcohol use, and 3) pitfalls of these opening discussions.

##### Establishing Rapport

In our focus groups, local professionals agreed that the initial greeting between patient and provider sets the tone for the entire BNI and should therefore be conducted in a way that maximizes comfort on the patient’s behalf. Given the importance of the initial greeting in our setting, all team members agreed that sitting if possible, introducing oneself, and inquiring if the patient’s health concerns have been addressed should define the initial interaction between patient and provider. Step one of our adapted BNI protocol therefore begins with customary greetings.

##### Introducing the Topic of Alcohol

In our focus groups, it was agreed the topic of alcohol should only be raised after culturally appropriate introductions and greetings have been exchanged and the patient’s primary health concerns, such as pain or hunger, have been addressed. Addressing patient concerns was therefore included in step one of our adapted BNI protocol, following customary greetings and preceding the introduction of alcohol use. Furthermore, before directing the conversation towards alcohol the provider should ask the patient for permission to discuss alcohol, to show respect. Step one therefore requires patient permission prior to opening a discussion about alcohol use.

##### Pitfalls of Opening Discussion

Local professionals in our focus groups emphasized the importance of the characteristics of the person who conducts the BNI. Given the influence of one’s cultural environment, demographic factors such as age and gender are very important and need to be considered due to the sensitive or stigmatizing topics to be discussed.(Lakes et al., 2006; Field et al., 2010; Johnson et al., 2010; McNeely et al., 2018). Our intervention is specifically geared for patients at least 18 years of age or older, so the age of both patient and interviewer were crucial to consider as well. For example, according to our focus groups, participants thought it could be inappropriate and perhaps even counterproductive given the cultural context and norms to have a young male administer the BNI to an older male. Local professionals agreed and recommended that experienced female nurses, typical of Tanzania’s nursing population, would elicit the most productive feedback from patients. Again in the cultural context, it was recommended that older female nurses would be perceived more positively. As an example, when speaking to older female nurses, patients are perceived to feel more comfortable discussing alcohol because they are speaking to an elder. Patients also feel more inclined to listen to an elder female’s advice as they may feel as though they are speaking to their mother or someone similarly experienced. Consequently, our team agreed that delivery of our adapted BNI in the Tanzanian setting should be conducted by an experienced female nurse when possible.

Although older female nurses are ideal for administering the BNI in our setting, our focus groups also recognized the importance of ensuring that a male nurse is always available to deliver the BNI, also to accommodate cultural norms and the context. A particular and highly expected scenario was described pertaining to religion and cultural expectations with respect to gender. The local clinicians in our focus group agreed that Muslim male patients for example, may not be comfortable discussing alcohol with female providers because of their religious norms. Given Tanzania’s large Muslim population, our team agreed that having at least one male nurse available to deliver BNIs was a necessary accommodation to ensure that any patient in the Tanzanian setting had the opportunity to benefit from our intervention.

#### 2: Provide Feedback

The second step of the model BNI protocol reviews the patient’s drinking patterns and makes a connection between the patient’s alcohol use and their visit to the ED. Cultural adaptation of step two focused on 1) alcohol use patterns at the Tanzanian versus the global level, 2) use of the Alcohol Use Disorder Identification Test (AUDIT) as a marker of normative behavior and whether it is associated with increased risk in the Tanzanian setting, 3) culturally and linguistically appropriate Swahili words to describe alcohol use, alcohol amounts, and alcohol use disorders, and 4) the most common complications of alcohol use in our setting.

##### Population Level Alcohol Use Patterns

To help patients compare their alcohol use to societal norms, our focus groups thought it important to relate a patient’s alcohol use behavior to alcohol use patterns at the population level. Overall, our focus groups discussed how the population presenting to our health system is mostly a local population who are somewhat insulated and as such, international guidelines are less relatable to the typical patient than national or regional guidelines. Our team agreed that offering patients a description of alcohol norms from a regional perspective would allow patients to better contextualize their own alcohol related behavior. However, our team suggested allowing the discussion around alcohol norms to remain malleable. The conversation should begin with regional alcohol norms, but can shift to national or international norms if the interventionist feels these norms are more in line with the patient’s own alcohol related behavior. As an additional means of helping patients contextualize and quantify their alcohol use within the regional population, we created a visual guide to standard drinks (Figure 2) that can be shared during the discussion of population level alcohol use patterns.

**Figure 2:**
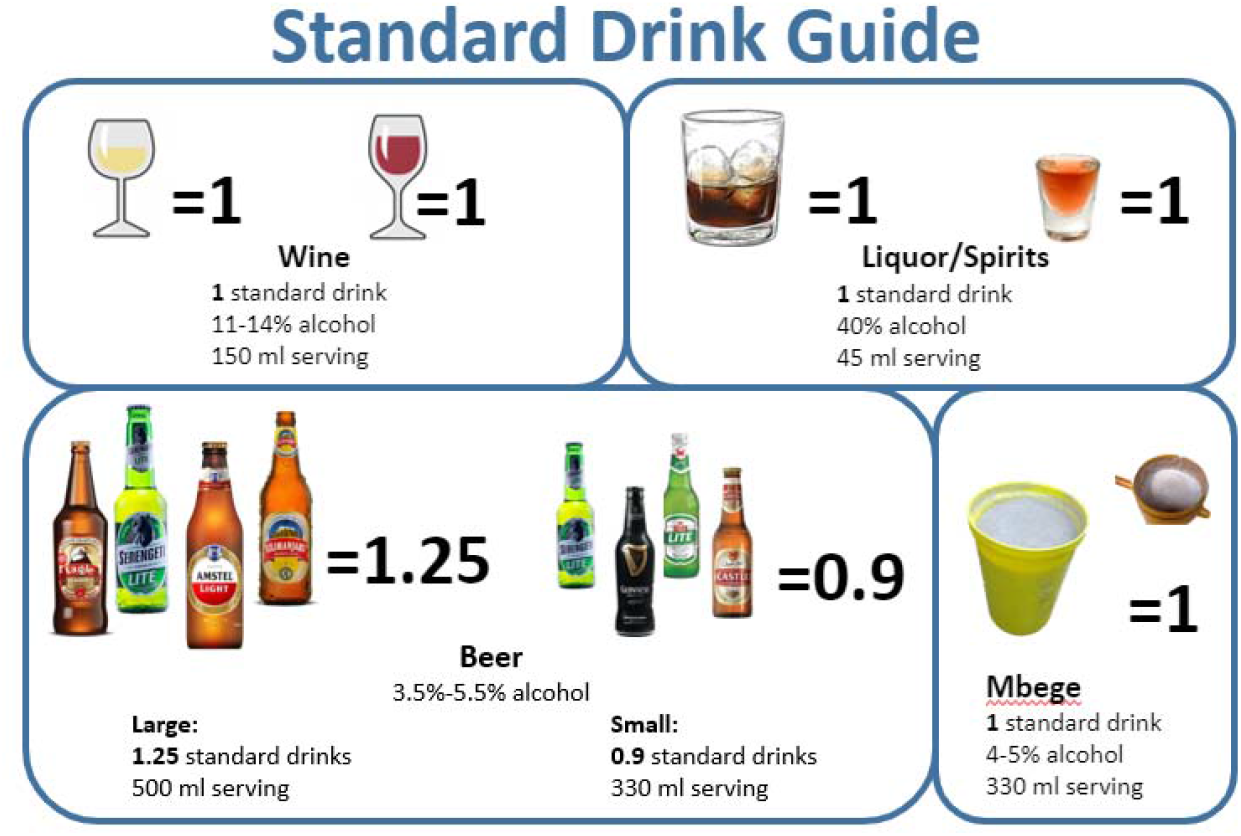
Our Standard Drink Guide for the Tanzanian Setting

##### Administration and Explanation of the AUDIT

Our team has previously conducted multiple validations of the AUDIT in Tanzania’s injury patient population to ensure both content and clinical validation. Our research team was also trained on how to make AUDIT results understandable to patients. Consequently, our team decided the AUDIT was the best way to screen ED patients for hazardous and harmful alcohol consumption. Step two of our adapted BNI protocol therefore offers the patient feedback regarding their AUDIT score. To facilitate discussion regarding the AUDIT, our team used the Range, Anecdote, Normal, Give, Elicit (RANGE) model to assist nurses in discussing AUDIT results. In the RANGE model the interventionist states the range of possible AUDIT scores, provides an anecdote exemplifying the AUDIT’s validity, states the range of scores corresponding to normal alcohol use, reveals the patient’s AUDIT score, and elicits feedback from the patient by asking what he or she thinks of their score.

##### Linguistic Adaptations

In our focus groups, linguistic and culturally appropriate non-judgemental words were chosen and enforced by our bilingual team performing the intervention (Table 3). We were very careful not to include any Swahili translations of English words regarding alcohol which might carry negative connotations.

**Table 3.**
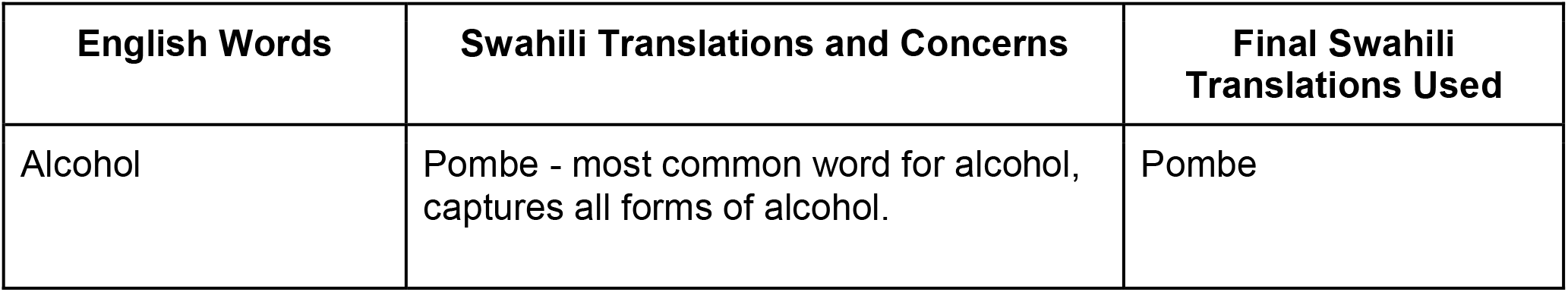
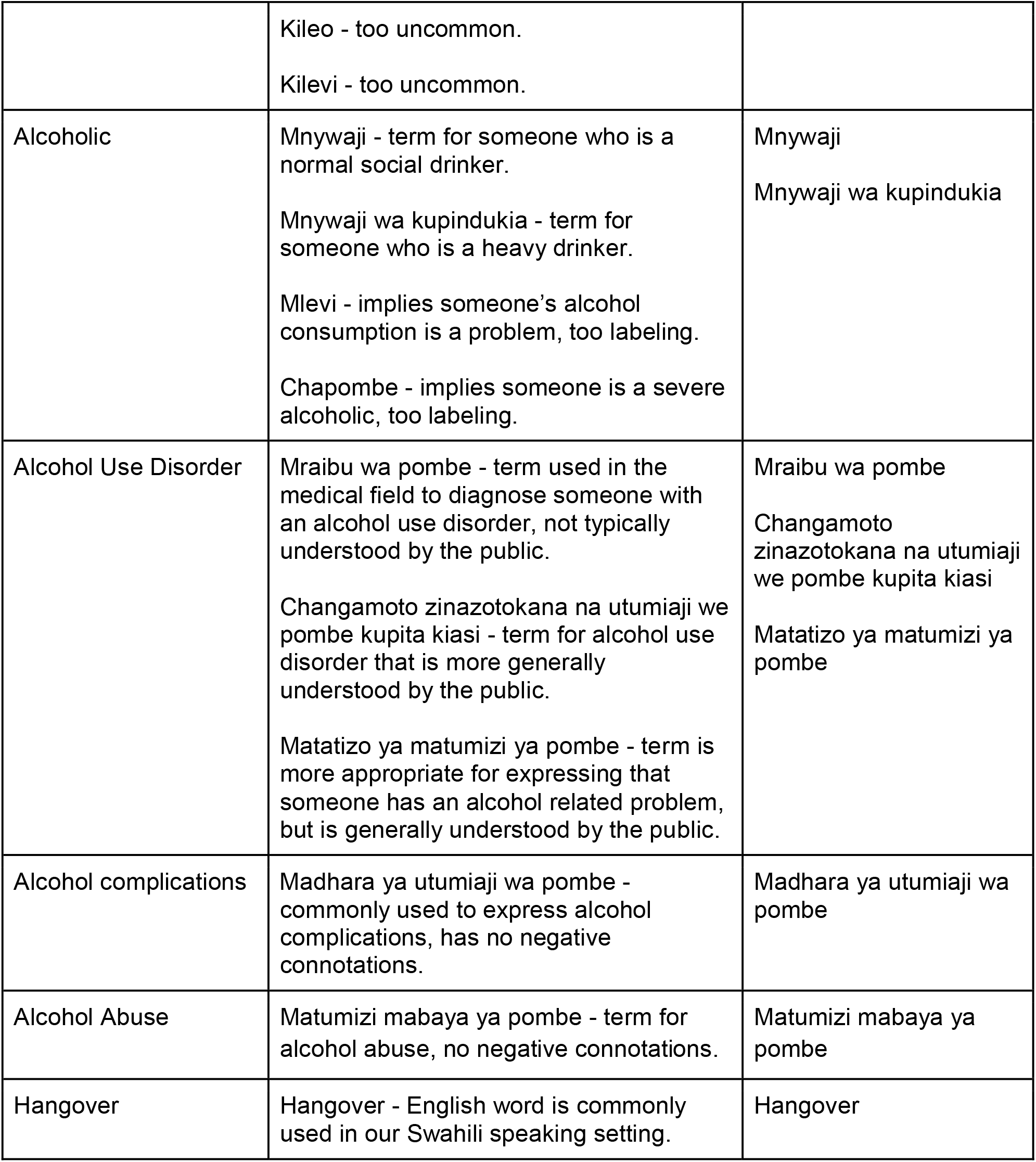
Swahili Translations of English Alcohol Words

##### Common Complications of Harmful Alcohol Use

Local professionals in our focus groups agreed that money, relationships, work/employment, and health are the most common areas of life in which alcohol creates complications among patients in our population. In the last portion of step two in our adapted BNI protocol, the interventionist shares these common alcohol related consequences with the patient. The purpose of sharing these consequences is to encourage the patient to consider and discuss how alcohol adversely impacts his or her own life.

#### 3: Enhance Motivation

The third step of the model BNI protocol assesses a patient’s readiness to change their alcohol related behavior and incorporates open ended questions about their readiness to change as well as reflective listening. Cultural adaptation of step three focused on 1) the development of a readiness to change ruler appropriate to our setting, 2) making a link between alcohol-related consequences and a patient’s alcohol use, 3) making a link between a patient’s personal problems and their alcohol use, and 4) using motivational techniques to encourage behavior change.

##### Developing a Readiness to Change Ruler

All members of our focus group decided that a 0-100 scale ranging from “not at all ready” to “extremely ready” was appropriate for assessing readiness to change in our setting. Initially, some of our external professionals opposed a 0-100 scale given the nature of the question. A self-evaluation of readiness to change requires significant thought and reflection. A 0-100 scale presents too large a range of possibilities to choose from which may hinder or stall the goal of assessing readiness. However, local members expressed strong support for a 0-100 scale because numerical expressions in terms of percentages are more common in Tanzanian society. Thus, a 0-100 scale would make it easier for patients to assess readiness to change.

External professionals in our focus groups also noted that the goal of assessing readiness to change is to understand why a patient thinks he or she needs to change. Consequently, we added the following question to supplement the readiness to change ruler: “why did you not pick a lower number?” Such a question encourages the patient to defend their reasons for being ready to change, rather than the contrary, “why did you not pick a higher number?” which highlights barriers to change.

##### Establishing a Link Between Alcohol Use, its Consequences, and Personal Problems

Focus groups agreed that the link between alcohol use and alcohol related consequences, such as injuries, is not obvious to many patients in our setting. Moreover, our team concluded that there is a general lack of education among most patients in our setting regarding safe drinking limits. Step three of our adapted BNI protocol therefore establishes a link between harmful alcohol use and alcohol related consequences, and a subsequent link between harmful alcohol use and the patient’s personal problems resulting from hazardous drinking behaviors. Members of our focus groups emphasized the importance of establishing these two links. If a patient does not understand the connection between their alcohol use and their personal problems, then he or she is unlikely to be receptive towards step 4 of our adapted BNI protocol which prompts behavior change.

##### Encouraging Behavior Change

Focus groups revealed that care in the Tanzanian setting if often provided in a hierarchical manner in which patients are given instructions in the form of commands (ie. “do this” or “do not do this”). Encouraging, rather than telling, patients to change their behavior is uncommon. Consequently, step three of our adapted BNI protocol ends with the interventionist summarizing the patient’s responses from this step using motivational techniques outlined in the model BNI protocol. These techniques include expressing empathy towards the patient, developing discrepancy between the patient’s alcohol use behavior and good health, “rolling with” a patient’s resistance to behavior change so as to avoid confrontation and elicit productive feedback, and supporting the patient’s self-efficacy and confidence to build motivation.

#### 4: Negotiation and Advice

The fourth step of the model BNI protocol creates a plan for the patient to reduce harm resulting from alcohol use, offers advice to the patient regarding their alcohol use, and formalizes an agreement between the patient and the interviewer. Cultural adaptation of step four focused on 1) developing an array of patient options for reducing alcohol related harm, 2) discussing the most effective way to offer advice to a patient, 3) discussing the most practical way to formalize an agreement with a patient, and 4) summarizing the results of the intervention.

##### Reducing Alcohol Related Harm

Our focus groups concluded that the following options for reducing alcohol-related harm are most appropriate for patients in our setting: manage drinking, eliminate drinking from your life, never drink and drive, limit drinking, and seek help. In step four of our adapted BNI protocol, these options are presented to a patient who is then asked to choose which option they would prefer to use. It is important to note that each of these options may not apply to each patient. Thus, options should be modified and presented to individuals at the discretion of the interventionist.

##### Offering Advice

Members of our focus groups agreed asking patients for permission to offer advice is not necessary because the intervention implies that professional advice will be given. However, we included asking for permission to give advice in step four of our adapted BNI protocol as a way of keeping the patient comfortable and in control of the intervention.

##### Formalizing an Agreement

To reinforce a patient’s commitment to behavior change, all focus group members agreed that a verbal agreement was most appropriate as many patients in our Tanzanian setting have low education and as such relatively limited writing capacity and a low literacy rate. Focus group members also agreed that asking patients to identify a potential supporter of change should be part of the agreement process to make patients feel like they do not have to confront behavior change alone. However, identifying a personal supporter of change is entirely optional as some patients may feel uncomfortable sharing private information with family or friends.

##### Summarizing Results of the BNI

The final element of step four in our adapted BNI protocol is to present a summary of the intervention that emphasizes the patient’s statements, strengths, and agreement. All members of our focus group agreed that the summary should end by giving control back to the patient. Following the summary, we therefore added the following question: “Are there any additional statements you would like to add?” By asking this final question, the patient has the opportunity to end the intervention with their own concluding remarks.

## DISCUSSION

Through a multiphase iterative process driven by input from local healthcare professionals and supplemented with guidance from external professionals, we culturally adapted a BNI to the Tanzanian setting, culture, and language. To our knowledge this is the first AUD intervention to be adapted specifically for acute injury patients in an LMIC population.

### The Need for Cultural Adaptation

The prevalence of alcohol abuse in the Kilimanjaro region of Tanzania is among the highest in the country, exceeding values representative of other regions as well as the entire nation (Mitsunaga et al., 2008; Francis et al., 2015; Mbatia et al., 2009; WHO, 2018). The even larger prevalence of self-reported or clinician-detected alcohol use among injury patients seeking emergency care highlights the need for evidence-based AUD interventions that can be implemented in Tanzanian EDs (Mundenga et al., 2019; Staton et al., 2018). To be effective, such interventions require adaptation to the local cultural context. Our study demonstrates that cultural adaptation of a substance use intervention can encompass a broad range of changes to the original intervention protocol. These changes may determine who administers the intervention, how a patient is greeted, how the topic of substance abuse is raised, how a patient is informed of their harmful substance use, how graphics are visualized within the intervention protocol, how behavior change is motivated, and which behavior changes are encouraged. While such changes may not be required in every unique cultural setting, the results of our study provide a framework that may help construct cultural adaptation procedures for substance use interventions in other LMICs.

Cultural adaptations have been applied to BNIs for non-Tanzanian populations at increased risk of harmful substance use, and have indicated high participant satisfaction (Torres et al., 2020; Tsarouk et al., 2007). In addition, culturally adapted substance use interventions have proven to be more effective than their non-adapted counterpart in reducing harmful behavior in a multitude of settings including both high-income countries and LMICs (Hecht et al., 2018; Papas et al., 2011; Papas et al., 2020). In Tanzania, however, there are no culturally sensitive alcohol use interventions that have been systematically developed and established across healthcare facilities (Meier et al., 2020). Our PPKAY tool is the first substance use intervention to be culturally adapted for administration to Tanzanian patients who test positive for harmful alcohol consumption. The numerous changes deemed necessary to make the original intervention protocol applicable to Tanzanian patients highlights the need for rigorous cultural adaptation of interventions developed outside of LMICs, but applied to LMIC populations.

### The Cultural Adaptation Process

Cultural adaptation refers to modifications of an evidence-based intervention that render the intervention more compatible with the cultural, social, and linguistic behaviors of a target population (Burlew et al., 2013). The adaptation process therefore relies on community engagement to support incorporation of these societal norms into the adapted intervention. Notably, a community advisory board (CAB) is commonly used throughout the iterative adaptation process to ensure appropriate cultural translation of a substance use intervention without compromise of the intervention’s core effective components (Whitesell et al., 2019; Field et al., 2015; Field et al., 2019; Hutton et al., 2019). In the Tanzanian setting, the use of a CAB alone to guide adaptation of our intervention was challenging due to limited knowledge among the target population regarding safe alcohol use behaviors, AUD interventions, and local AUD treatment options (Meier et al., 2020; Staton et al., 2018). Consequently, our approach to adaptation included input from a CAB, but relied more heavily on guidance from local healthcare professionals with extensive experience in substance use counseling. In addition, we sought input from external professionals with extensive substance use research experience to maintain fidelity of the original intervention protocol throughout the adaptation process. This approach is not entirely unique as it has been used for intervention adaptation in high-income settings (Whitty et al., 2016; Ornelas et al., 2015). However, our study shows that the collective knowledge of community members, local healthcare professionals, and external professionals creates a rich environment for cultural adaptation of substance use interventions in LMICs, even in settings where community-based participatory research is less common.

### Next Steps

Our PPKAY intervention was designed with the entirety of our target population in mind. In its current form, our BNI seeks to reduce harmful alcohol use for any injury patient with an AUD presenting to KCMC in Moshi, Tanzania. However, the Kilimanjaro region of Tanzania is incredibly diverse with regard to age, education, income, religion, tribe, and other sociodemographic factors. Intragroup diversity of this kind may create differences in intervention efficacy between subgroups (Burlew et al., 2013). As we implement our adapted BNI in the Tanzanian setting, it is important that we compare the efficacy of our intervention across these subgroups so that we may continue tailoring our BNI to individuals with specific backgrounds. Furthermore, there is substantial stigma across Tanzanian communities in the Kilimanjaro region towards individuals who exhibit harmful alcohol use (Meier et al., 2020; El-Gabri et al., 2020; Griffin et al. 2020; Staton et al., 2020). External and internalized stigma may prevent patients from engaging in conversations elicited by our BNI. It is therefore important that we ensure our intervention can create meaningful behavior change among those who feel stigmatized.

## CONCLUSION

We took a BNI originally developed to lower the risk of alcohol use among ED patients in the United States, and adapted it through a multiphase iterative process to create an AUD intervention applicable to ED patients in the Tanzanian setting. Our adapted tool, the PPKAY, is a 15-minute brief intervention that uses motivational interviewing techniques to inspire behavior change among injury patients who engage in harmful alcohol use. Our study outlines the specific steps taken to make the cultural adaptations of this BNI. Our study also demonstrates an approach for how cultural adaptation of substance use interventions can be made in low-income settings as well as settings where community knowledge surrounding substance abuse is limited. Given the scarcity of research on alcohol use in Tanzania and similar settings, we encourage others to explore the cultural adaptation and evaluation of interventions to reach populations in need. Meanwhile, we will test this adapted BNI at KCMC for efficacy and report the subsequent results in a future study.

## Data Availability

N/A

## Acknowledgements

To the KCMC Emergency Department research team, we thank you for your hard work, dedication and perseverance to improve the lives of your patients. Your dedication to your community is why projects like these happen.

